# Statin intolerance – Prevalence and practical management in a specialized lipid clinic

**DOI:** 10.1101/2022.05.05.22274716

**Authors:** Yannick Turdo, Tinh-Hai Collet

## Abstract

**Aims:** Statins are recommended as first-line treatment to prevent cardiovascular disease, but high rates of adverse events and subsequent discontinuation can prevent effective treatment in some patients. We aimed to describe the intensity, nature and frequency of adverse effects caused by statins and other lipid-lowering drugs, and to assess their association with clinical predictors and treatment strategies.

**Methods:** This is a retrospective study of 121 consecutive unselected patients followed at a specialized lipid referral clinic in Lausanne University Hospital, Switzerland, in 2018 whose clinical data spanned 2016 until 2019.

**Results:** Adverse effects were reported by 58% of patients, causing musculoskeletal (72%), neurologic (40%), gastrointestinal (25%) and other (25%) symptoms. Gastrointestinal symptoms affected 35% of women, but only 12% of men (p=0.06). Statins caused more adverse effects than other drugs, with atorvastatin (63%) and simvastatin (64%) achieving the lowest rates, pravastatin (87%) and fluvastatin (100%) the highest. Fenofibrate was associated with significantly less frequent adverse effects than statins (22% vs. 57%, p=0.006). Adaptation and termination of the treatment led to 86% and 88% lower intensity of symptoms, respectively.

**Conclusion:** While the predominance of musculoskeletal symptoms caused by lipid-lowering drugs is known, symptoms affecting other organ systems should not be ignored. Statins were the lipid-lowering drug class with the highest rates of adverse effects. To maintain compliance and cardiovascular prevention, treatment strategies such as a change of dosage, frequency of administration, daily timing, or switching active substance can contribute to better tolerance of statin treatment. Further investigation is needed to establish specific treatment strategies for individual patient profiles.

## Introduction

Statins inhibit the 3-hydroxy-3-methyl-glutaryl-coenzyme A reductase enzyme found in the liver, thus reducing endogenous production of cholesterol. This drug class has been the recommended first-line treatment to prevent cardiovascular diseases (CVD) for decades.^1,2^ Due to the high prevalence and clinical burden of CVD, a large number of patients are prescribed statins worldwide. This has made statins one of the best-selling drug classes, with currently 6 active substances available in Switzerland: pravastatin, simvastatin, fluvastatin, atorvastatin, rosuvastatin and pitavastatin.

Unfortunately, adverse effects are frequently reported by patients on statins. Muscle pain and other musculoskeletal symptoms are the most common complaints, affecting up to 17% of statin users,^3^ grouped in the emerging entity statin-associated muscle symptoms (SAMS). Less frequently reported effects include gastrointestinal, neurological and dermatologic symptoms, increased risk of type 2 diabetes, reduced renal function and abnormal liver function tests.^4^

Due to these adverse effects, drug management can become difficult for patients and their healthcare providers. Treatment often needs adjustments to avoid, or at least decrease, the reported adverse effects. Options used by clinicians include lowering the dosage, temporary discontinuation (also known as drug holiday), or switching to a different (statin or non-statin) lipid-lowering drug.^5,6^

Adverse effects have a significant impact on quality of life, sometimes leading to lower adherence to the treatment or failure to follow-up.^4,7^ Patients develop statin intolerance, i.e. the inability to tolerate the required dose of statin to sufficiently reduce their risk of CVD. This may cause the discontinuation of important medication, thus putting patients at increased risk of future CVD.^8^ Statins have fallen into disrepute over the years and patients often look for alternative medication. Medical management requires substantial time and interdisciplinary management to ensure that patients do not definitely discontinue their treatment.^1^

Interestingly, statin intolerance is reported with different frequency according to sex, age groups and comorbidities (e.g. hypothyroidism, diabetes and muscle disorders). Additionally, the different metabolic pathways of individual statins (cytochromes), and the hydrophilic vs lipophilic nature of the active substance lead to varying profiles of drug interaction.^4^ It is important for clinicians to use personalized strategies to adapt the lipid-lowering drug regimen to decrease the rate of adverse events and increase drug adherence. Algorithms and recommendations which aim to facilitate management of statin intolerance have been proposed.^9–11^ Knowing further factors associated with statin intolerance may help to improve on such strategies. The literature on statin intolerance is still limited and its prevalence in Switzerland is currently unknown.

In a specialized lipid clinic at a Swiss University Hospital, we retrospectively reviewed the electronic medical records of 121 patients to explore the intensity and nature of statin associated adverse effects, and to assess the association of statin intolerance with clinical predictors, such as demographics, comorbidities or other medication. Furthermore, we explored the different strategies used by clinicians when facing statin intolerance. By evaluating and comparing the rate of success among them, we aim to improve clinical management and cardiovascular prevention coverage.

## Materials and Methods

### Study design

This retrospective study included a random subset of the patients treated at the tertiary lipid clinic of the Service of Endocrinology, Diabetes and Metabolism at Lausanne University Hospital (CHUV) in Lausanne, Switzerland. The local ethics committee (CER-VD, Lausanne, Switzerland) approved the study protocol and waived the requirement for individual informed consent of this anonymous retrospective review of electronic medical records.

### Data collection

Data was abstracted from the electronic medical records and stored in a secured database. This was done by a single investigator (Y.T.), thus eliminating potential discrepancies caused by multiple reviewers. An unselected consecutive subset of the 376 unique patients visiting the lipid clinic during 2018 was drawn, due to time and resource constraints. We collected data from all clinic visits of the 133 drawn patients based on archived medical letters sent to their general practitioners, thus spanning the period between November 29, 2016 and July 8, 2019. Of note, 12 patients had no archived medical letters referencing their clinic visits during this period, therefore the final data analyses were performed on the clinical data of 121 patients (**Figure 1**).

**Figure 1.**
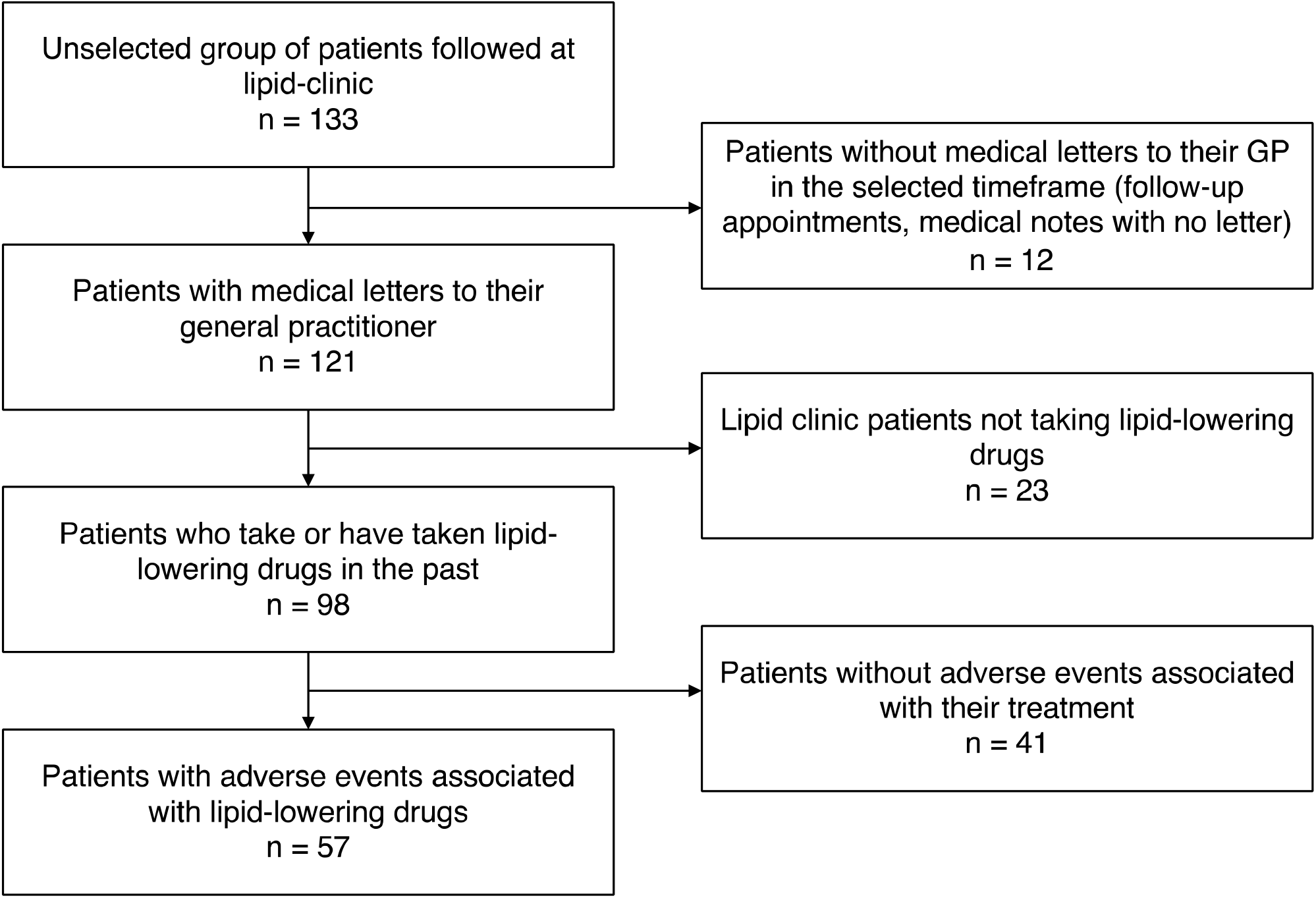
Flowchart of patients’ data used in the study An unselected consecutive subset of the 376 unique patients visiting the lipid clinic during 2018 was drawn. We collected data from all clinic visits of the 133 patients based on archived medical letters sent to their general practitioners (GP), thus spanning the period between November 29, 2016 and July 8, 2019. Of note, 12 patients had no archived medical letters about their clinic visits during this period, therefore the final data analyses were performed on the clinical data of 121 patients.

Demographics, medical history, medication, laboratory results and adverse effects attributed to lipid-lowering drugs were extracted from electronic medical records. Data on medication, laboratory results and adverse effects were collected from each individual patient visit, while demographics and medical history were recorded only once per patient. When available, we recorded data on adverse effects attributed to lipid-lowering drugs, which took place before the patient’s first visit.

In addition to baseline characteristics, we abstracted data on cardiovascular, musculoskeletal, neurologic and metabolic disorders (e.g. hereditary muscle disorders, diabetes, obesity defined as body mass index [BMI] ≥ 30 kg/m^2^, vitamin D deficiency, thyroid, liver, kidney status), physical activity, medication, laboratory results. As physical activity was not recorded in a standard fashion in electronic medical records, it was rated by one investigator (Y.T.) into one of 2 categories: active and inactive, based on the information provided by the clinicians.

Symptoms associated with lipid-lowering drugs were recorded individually and grouped into 4 organ systems: musculoskeletal, neurologic, gastrointestinal, and others.^4^ These adverse effects were recorded during the interval between visits at our clinic, but also historically before the first visit (for example when mentioned by the primary care physician). In addition, based on the patient’s complaints, the intensity of the symptoms was rated by the only reviewer (Y.T.) into one of 3 categories: low, medium and high, when possible. The physician’s strategy in the face of adverse events was recorded as continuation, termination or adaptation of treatment, including change of dosage, frequency or medication.

### Statistical analyses

Continuous variables are reported as means ± standard deviations (SD), while frequencies (percentage) are used for categorical variables. For comparison of categorical values, Pearson’s Chi^2^ test was used, except for fewer than 5 observations in a single category in which case Fisher’s exact test was employed. P values < 0.05 were considered statistically significant. Statistical analyses were performed using STATA software package (version 15.1, College Station, TX, USA).

## Results

### Baseline Characteristics

Among the 121 patients, the mean age was 55.7 ± 12.5 years, the mean BMI was 27.0 ± 4.4 kg/m^2^ and 57 (47%) were women (**Table 1**, *detailed in Table S1*). The prevalence of CVD was 52%, hypertension 41%, current smoking 29%, diabetes 16%, obesity 24% and chronic kidney disease 9%. At the first clinic visit, 63 (52%) of patients used statins and 40 (33%) used a non-statin lipid-lowering drug, such as ezetimibe and fenofibrate. In total, 98 (81%) patients took a lipid-lowering drug of any kind at least once in their lifetime before their first clinic visit.

**Table 1.**
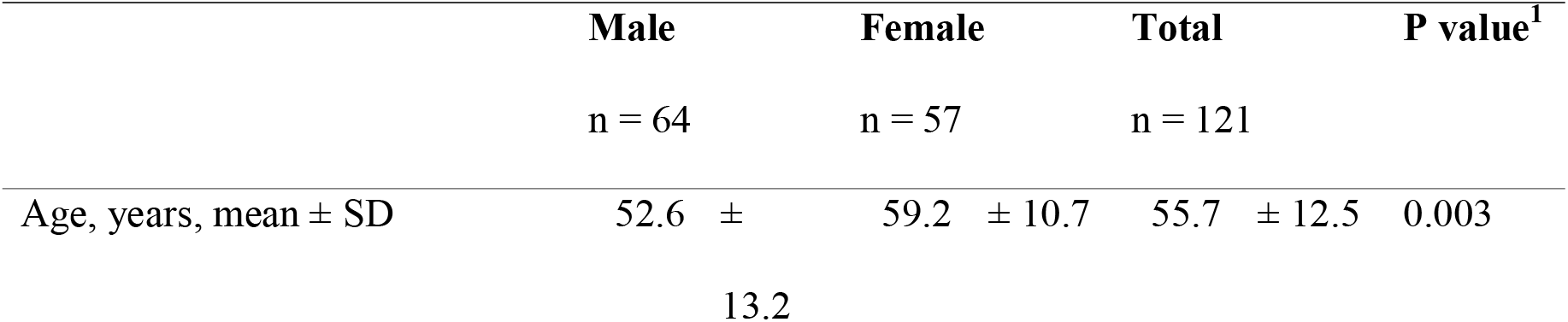
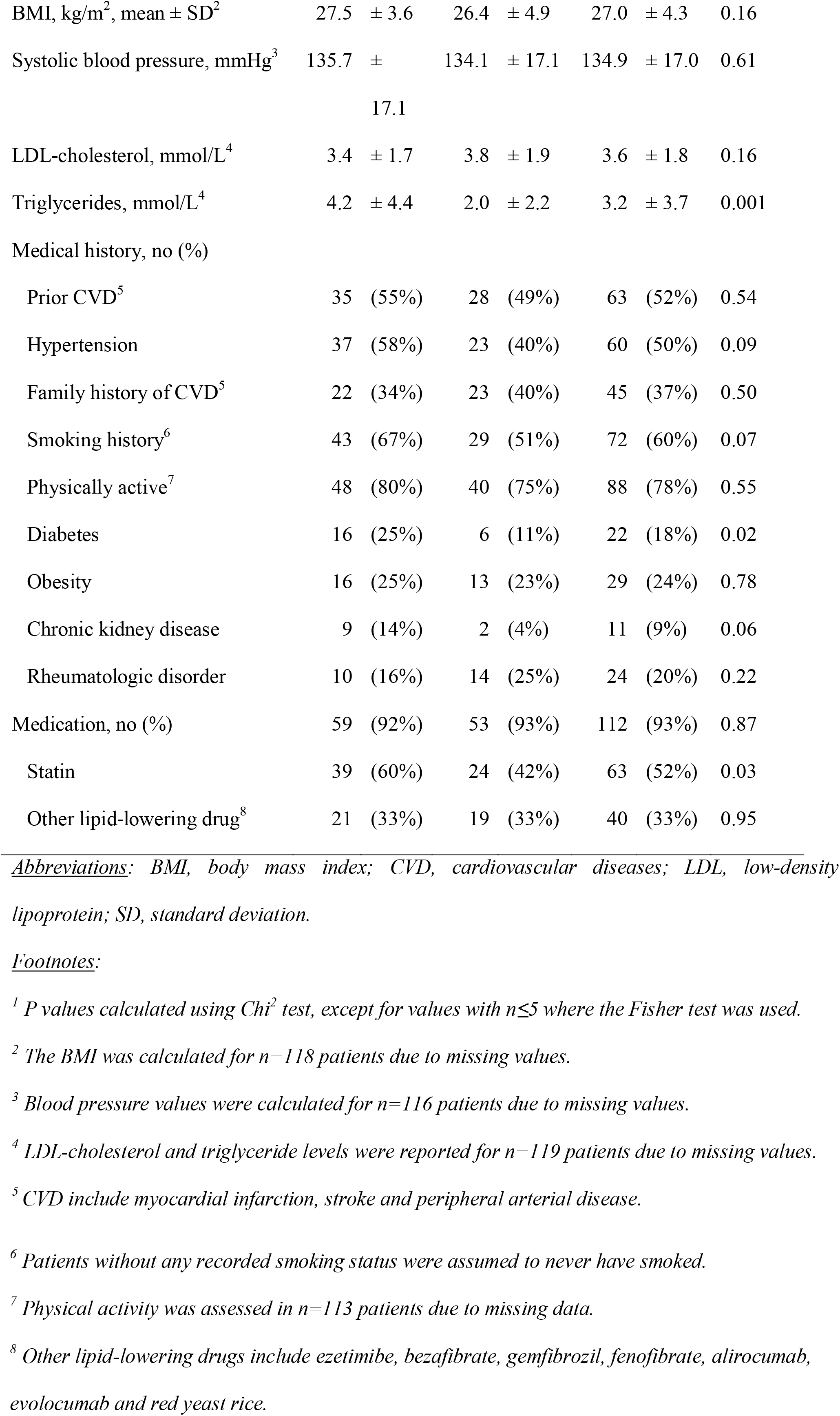
Clinical characteristics of 121 patients followed at lipid clinic

### Lipid-lowering drug associated adverse events

Among the 98 patients who took lipid-lowering drugs (**Figure 1**), 57 (58%) reported adverse effects, which occurred during medical care at the lipid clinic or before (e.g. under the primary care physician’s care). The majority of patients (n=34, 60%) reported symptoms of a single organ system, while 9 patients (16%) experienced adverse events spanning 3 or more of the 4 organ system groups (**Table 2**).

**Table 2.** Lipid lowering drug associated adverse effects by organ system

|  | Male | Female | Total | P value<sup>1</sup> |
| --- | --- | --- | --- | --- |
| Affected organ systems | n = 26 | n = 31 | n = 57 | 0.13 |
| **Musculoskeletal** | **18 (69%)** | **23 (74%)** | **41 (72%)** | **0.68** |
| Myalgia | 14 (54%) | 20 (65%) | 34 (60%) |  |
| Arthralgia | 3 (12%) | 5 (16%) | 8 (14%) |  |
| Cramps | 3 (12%) | 4 (13%) | 7 (12%) |  |
| Rhabdomyolysis | 1 (4%) | 0 (0%) | 1 (2%) |  |
| **Neurologic** | **11 (42%)** | **12 (39%)** | **23 (40%)** | **0.78** |
| Fatigue | 4 (15%) | 7 (23%) | 11 (19%) |  |
| Headache | 2 (8%) | 6 (19%) | 8 (14%) |  |
| Dizziness | 4 (15%) | 1 (3%) | 5 (9%) |  |
| Insomnia | 0 (0%) | 3 (10%) | 3 (5%) |  |
| Confusion | 1 (4%) | 1 (3%) | 2 (4%) |  |
| Memory Loss | 1 (4%) | 1 (3%) | 2 (4%) |  |
| Depression | 2 (8%) | 0 (0%) | 2 (4%) |  |
| Anxiety | 1 (4%) | 0 (0%) | 1 (2%) |  |
| Paraesthesia | 0 (0%) | 1 (3%) | 1 (2%) |  |
| **Gastrointestinal** | **3 (12%)** | **11 (35%)** | **14 (25%)** | **0.062** |
| Nausea | 3 (12%) | 5 (16%) | 8 (14%) |  |
| Abdominal pain | 2 (8%) | 4 (13%) | 6 (11%) |  |
| Abdominal bloating | 1 (4%) | 3 (10%) | 4 (7%) |  |
| Vomiting | 2 (8%) | 1 (3%) | 3 (5%) |  |
| Diarrhoea | 1 (4%) | 2 (6%) | 3 (5%) |  |
| Constipation | 0 (0%) | 4 (13%) | 4 (7%) |  |
| Acid Reflux | 0 (0%) | 1 (3%) | 1 (2%) |  |
| **Others** | **3 (12%)** | **11 (35%)** | **14 (25%)** | **0.062** |
| Rash | 1 (4%) | 1 (3%) | 2 (4%) |  |
| Elevated liver function tests | 0 (0%) | 2 (6%) | 2 (4%) |  |
| Dry Mouth | 0 (0%) | 2 (6%) | 2 (4%) |  |
| Heart palpitations | 0 (0%) | 1 (3%) | 1 (2%) |  |
| Oedema | 0 (0%) | 1 (3%) | 1 (2%) |  |
| Other symptoms<sup>2</sup> | 2 (8%) | 6 (19%) | 8 (14%) |  |

<sup>1</sup> *P values calculated using Chi<sup>2</sup> test, except for single values <5 where the Fisher test was used. The comparisons for specific symptoms were all non-significant and are thus not reported for legibility.*
<sup>2</sup> *Other symptoms included malaise, dyspnoea, itches, aphthae, hot flushes, loss of libido*

Musculoskeletal symptoms were by far the most common among the 4 organ systems (p<0.001), being experienced by 41 out of the 57 (i.e. 72%) patients reporting lipid-lowering drug associated adverse effects (**Table 2**). Among individual symptoms, myalgia was the most common (n=34, 60%), followed by arthralgia (n=8, 14%) and cramps (n=7, 12%). Only one case of rhabdomyolysis was recorded (2%). Among the other organ systems, general fatigue (n=11, 19%) was the most frequently reported, followed by nausea (n=8, 14%) and headaches (n=8, 14%).

In the comparison between sex, 31 women (54%) and 26 men (41%) experienced adverse effects, but there was no significant difference (p=0.13). Among the individual organ system groups, both women and men reported musculoskeletal symptoms (p=0.68) and neurologic symptoms (p=0.78) similarly, while more women (n=11, 35%) reported gastrointestinal symptoms than men (n=3, 12%) with borderline significance (p=0.062). The remaining symptoms (grouped in ‘Others’ in **Table 2**) also affected women (n=11, 35%) more often than men (n=3, 12%) with borderline significance (p=0.062).

Of the 24 patients suffering from rheumatological disorders, 11 (46%) experienced adverse effects, as did 6 (55%) of the 11 patients with chronic kidney disease, 2 (67%) of the 3 patients with liver disease and 5 (50%) of the 10 patients with thyroid disorders. These adverse effect rates did not differ from patients without these conditions (all p≥0.28).

### Comparison of adverse effects across individual lipid-lowering drugs

We explored the frequency of adverse effects between the different lipid-lowering drugs. Of the 98 patients on lipid-lowering drugs before or during the clinic visit, 87 (89%) took statins, 50 (51%) ezetimibe, and 32 (33%) other drugs (**Table 3**). Multiple patients took more than one lipid-lowering drug, sometimes at the same time. This suggests that for these patients, the cause of adverse events might be confounded and the rate reported might be too high for drugs frequently prescribed with others, most notably ezetimibe. In this study, 96% of patients who took ezetimibe also were on statins.

**Table 3.** Number of patients reporting adverse effects associated with specific lipid-lowering drugs

|  | Any<br>symptoms | Musculo-<br>skeletal | Neurologic | Gastro-<br>intestinal | Other | None | Total<sup>1</sup> |
| --- | --- | --- | --- | --- | --- | --- | --- |
| **Statins** | **50 (57%)** | **39 (45%)** | **20 (23%)** | **10 (11%)** | **13 (15%)** | **37 (43%)** | **87** |
| Atorvastatin | **38 (63%)** | 32 | 11 | 7 | 10 | **22 (37%)** | **60** |
| Fluvastatin | **3 (100%)** | 3 | 0 | 0 | 0 | **0 (0%)** | **3** |
| Pitavastatin | **12 (67%)** | 9 | 4 | 2 | 2 | **6 (33%)** | **18** |
| Pravastatin | **13 (87%)** | 10 | 5 | 1 | 3 | **2 (13%)** | **15** |
| Rosuvastatin | **28 (67%)** | 23 | 11 | 3 | 9 | **14 (33%)** | **42** |
| Simvastatin | **9 (64%)** | 8 | 2 | 1 | 2 | **5 (36%)** | **14** |
| **Ezetimibe** | **28 (56%)** | **21 (42%)** | **9 (18%)** | **6 (12%)** | **7 (14%)** | **22 (44%)** | **50** |
| **Fibrates** | **5 (25%)** | **4 (20%)** | **0 (0%)** | **1 (5%)** | **1 (5%)** | **15 (75%)** | **20** |
| Bezafibrate | **0 (0%)** | 0 | 0 | 0 | 0 | **1 (100%)** | **1** |
| Gemfibrozil | **2 (67%)** | 1 | 0 | 1 | 0 | **1 (33%)** | **3** |
| Fenofibrate | **4 (22%)** | 4 | 0 | 0 | 1 | **14 (78%)** | **18** |
| **PCSK9 inhibitor** | **5 (45%)** | **4 (36%)** | **4 (36%)** | **3 (27%)** | **1 (9%)** | **6 (54%)** | **11** |
| Alirocumab | **1 (25%)** | 1 | 1 | 1 | 0 | **3 (75%)** | **4** |
| Evolocumab | **4 (57%)** | 3 | 3 | 2 | 1 | **3 (43%)** | **7** |
| **Red yeast rice** | **3 (100%)** | **3 (100%)** | **1 (33%)** | **1 (33%)** | **1 (33%)** | **0 (0%)** | **3** |

<sup>1</sup> Total number of individual patients on these drug classes or individual drugs. Percentages are calculated based on the total number of patients using the drug (e.g. 50 (57%) of the 87 patients on statins reported adverse effects associated with statins). Individual patients could be on multiple different drugs and therefore be listed under multiple sections.

Of the 87 patients on statins, 50 patients (57%) reported adverse effects, which was a higher rate than for any other group of lipid lowering-drugs with more than 3 patients treated (p=0.06). The rate of adverse effects for individual statins was even higher, as patients who experience adverse effects typically switch to a different statin and then commonly experience similar adverse effects. The two most commonly used statins in our clinic were atorvastatin (60 patients) and rosuvastatin (42 patients). They showed a similar rate of adverse events (63% and 67%, respectively, p=0.73). Simvastatin (64%) and pitavastatin (67%) showed similar rates, while pravastatin recorded a higher rate of 87%, much higher than the lowest rate achieved by atorvastatin (p=0.12).

**Table 3** also presents the rates of adverse effects in non-statin lipid-lowering drugs: the most commonly used drug was ezetimibe with 56% rate of adverse effects, most commonly associated with a statin, which likely caused frequent misattribution of symptoms. Fenofibrate was associated with less frequent adverse effects at only 22%, which is a significant difference compared to statins (p=0.006). Red yeast rice, while not admissible for treatment in Switzerland, was taken by 3 patients, all of whom reported adverse effects.

Musculoskeletal symptoms were the most common being experienced by over 40% for most medication classes containing more than 4 patients, the only exception being fenofibrate with 22% (p=0.055). Fenofibrate did not cause any neurological or gastrointestinal symptoms while only one out of 18 patients reported a non-musculoskeletal symptom. The numbers of adverse effects in individual organ systems were too low for meaningful statistical comparisons.

### Treatment Strategies

Treatment strategies and their adjustments were recorded in 90 instances of lipid-lowering drug associated adverse effects. While 13 (14%) patients continued with the same medication and dosage, 33 (37%) patients terminated treatment altogether (**Table 4**). Finally, 44 (49%) patients adjusted their treatment by either changing the dosage (n=5, 6%), switching the active substance itself (n=35, 39%), or changing the daily timing or frequency of administration (n=4, 4%).

**Table 4.** Adverse event intensity after re-evaluation of treatment strategy^1^

|  | **Total** | Lower | Same | Higher |
| --- | --- | --- | --- | --- |
|  |  | intensity | intensity | intensity |
|  | **n = 90** | n = 70 | n = 19 | n = 1 |
| **Continuation** | **13 (14%)** | 3 (23%) | 10 (77%) | 0 (0%) |
| **Adaptation** | **44 (49%)** | 38 (86%) | 5 (11%) | 1 (2%) |
| Change of dosage | **5 (6%)** | 5 (100%) | 0 (0%) | 0 (0%) |
| Change of frequency | **4 (4%)** | 4 (100%) | 0 (0%) | 0 (0%) |
| Switch medication | **35 (39%)** | 29 (83%) | 5 (14%) | 1 (3%) |
| Statin | **27 (30%)** | 23 (85%) | 3 (11%) | 1 (4%) |
| Non-statin | **8 (9%)** | 6 (75%) | 2 (25%) | 0 (0%) |
| **Termination** | **33 (37%)** | 29 (88%) | 4 (12%) | 0 (0%) |

*Footnotes:* <sup>1</sup> Some individual patients were subject to multiple changes in treatment
*Legend:* Number of patients and the evolution of their symptoms after a continuation, adaptation to the strategy (ie. dosage, frequency, drug), or termination of their treatment.

Predictably, the intensity of the experienced symptoms did not diminish for most patients (77%) who continued with the same treatment and was lowered for a majority of patients (88%) who terminated their treatment altogether (p<0.001). Adaptation of the treatment also resulted in a significantly higher proportion of patients (86%) experiencing a lower intensity of adverse effects (p<0.001), compared to continuing the treatment (**Figure 2**). The intensity of lipid-lowering drug-associated adverse effects was thus lowered for both an adaptation of the treatment, such as a switch to another medication, and complete drug termination (p=0.68).

**Figure 2.**
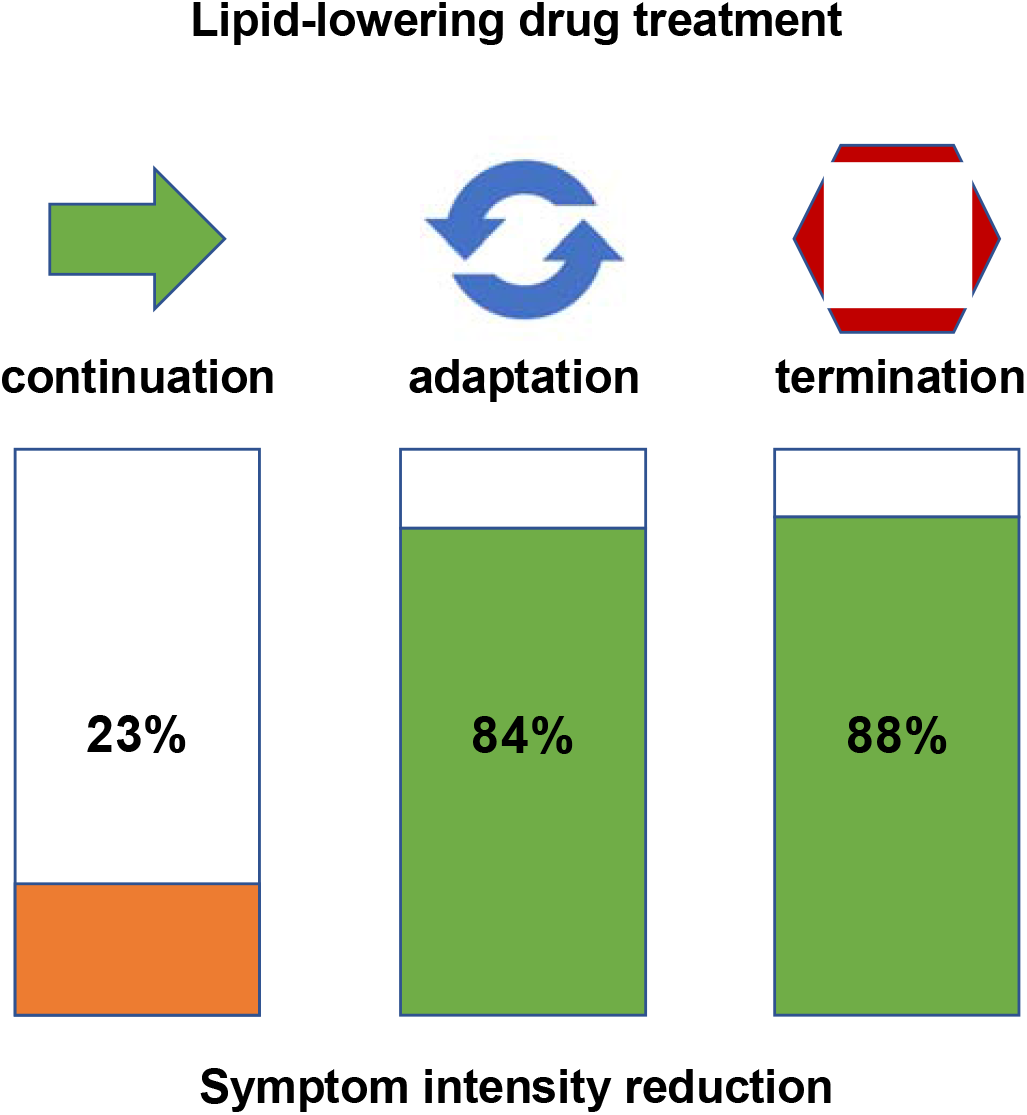
Symptom intensity reduction after adjustment of lipid-lowering drug treatment Adverse effects caused by lipid-lowering drug treatment require clinicians to choose between continuing, adapting or terminating treatment. Adjustments can range from a change of dosage, frequency of administration or daily timing, to switching the active substance. The rate of symptom intensity reduction could be successfully reduced with the adjustment strategy (86%) among 90 instances of strategy re-evaluation, compared to only 23% reduction with the continuation strategy and very close to the termination strategy (88%).

## Discussion

In our retrospective study of patients followed in a specialized lipid clinic, we found a clear predominance of musculoskeletal symptoms associated with the use of statins, as compared to adverse effects of other organs/systems. However, we found no significant difference in the frequency of adverse effects between sexes. When compared to other drug classes, statins caused more adverse effects than any other group (p=0.06), while fen of ibrate caused significantly less frequent adverse effects than statins (p=0.006).

This study did not find a significant difference in subsidence of adverse effect intensity between patients who discontinued their lipid-lowering drug treatment and patients who adjusted their treatment strategy or switched to a different lipid-lowering medication. This is strong encouragement for management strategies as they can provide the benefits of lipid-lowering treatment while not necessarily condemning the patient to higher intensity in adverse effects, than termination would.

As this study only included patients who were followed at a referral lipid clinic, the overall rate of statin associated adverse effects was expected to be higher than in studies which also include other types of patients and have found rates around 17%,^3^ compared to 57% in our study. The predominance of musculoskeletal symptoms does, however, mirror past studies on the topic.^3,4,12^ Furthermore, a predominance of musculoskeletal symptoms caused by other lipid-lowering drugs is also shown by this study, while previous literature on this topic is limited.

The comparison of lipid-lowering medication groups and their rates of adverse effects showed higher rates for statins than for any other group (p=0.06). While ezetimibe showed lower rates than individual statins, they were still higher than those of other agents. The very frequent association of ezetimibe with a statin might be the cause for these values as adverse events due to statins might be misattributed. When comparing individual statins which were taken by more than 10 patients, pravastatin showed higher rates (87%) than any other statin. Atorvastatin, pitavastatin, rosuvastatin and simvastatin all showed similar rates between 63% and 67%. This is not in line with previous studies, which have found atorvastatin to have the highest rates of adverse effects among statins (57%), with much lower rates for pravastatin (40%).^12^ It is indeed difficult to achieve a meaningful comparison in retrospective studies as the habits of clinicians to prefer one statin for first line treatment, as well as other factors, might interfere. One such factor might be a nocebo effect explored by previous studies.^13–15^ Fenofibrate caused significantly less frequent adverse effects than statins in this study (p=0.006).

A comparison between the frequency of adverse effects between sexes did not show any significant differences. Of note however, gastrointestinal symptoms were more often reported by women than men with borderline significance (p=0.062), while musculoskeletal (p=0.68) and neurologic symptoms (p=0.78) had no difference between sex, like in previous papers.^12^ A study including a larger cohort might find significant results.

When faced with patients suffering from lipid-lowering drug associated adverse effects, clinicians have multiple options to adjust treatment strategies in an aim to find a regime which is tolerable by the patient and sufficiently lowers the risk of CVD.^5,6^ While a change of dosage (n=5) or a change frequency or daily timing of administration (n=4) were occasionally used, this study found that most often clinicians switched the medication itself (n=35). This includes 27 changes to other statins and 8 changes to non-statin lipid-lowering drugs. Due to the small sample size, this study was unable to adequately compare different treatment strategies. We however found no significant difference in alleviation of symptoms between patients who had their treatment strategy adjusted and those who terminated their treatment altogether (p=0.68). This strongly suggests that it is worth for clinicians to persistently try alternative treatment strategies, such as a switch to a different drug, as even individual statins use different metabolic pathways and have different physical properties (hydrophilic vs lipophilic agents), making them more suited for different patients.^4^ Lipid-lowering drug associated adverse effects have been shown to increase the risk of recurrent CVD.^8^ The burden of CVD and the beneficial effects of lipid-lowering drugs are well documented.^1,2,16^ Clinicians are thus encouraged to be persistent and to find a treatment strategy acceptable for their patients with elevated cardiovascular risk, as rates of preventive care should be increased in most clinical settings, such as Switzerland.^17^

This retrospective study of patients’ records shares the limitations of retrospective studies, such as the clinical data having been recorded by multiple clinicians in a non-standardised way. *First*, although the single reviewer (Y.T.) assessed the clinical medical records in a consistent way, we cannot exclude that physical activity and the severity of symptoms were not recorded on a predefined scale, leaving them open to some interpretation. *Second*, we were unable to collect relevant data on the initial reason for referral to our lipid clinic, the indication and efficacy of treatments, the real life impact of adverse events, information on genetic predisposition, sufficient laboratory data for meaningful analysis, as well as the time interval between the introduction of treatment and the appearance of adverse events, from the available medical records. *Third*, the 23 patients who did not take lipid-lowering drugs may have been different from those taking statins, and recording of their symptoms was different from patients taking lipid-lowering drugs, making a case-control comparison impossible. *Fourth*, we did not record the dosage of medical treatment and can therefore not assess its impact on the frequency and intensity of adverse events. *Fifth*, all patients in this study were followed at a single specialized lipid clinic in Switzerland, which made data interpretation easier, as style of recording between clinicians did not vary as much as it would, presumably, between multiple clinics. This context however limits the generalizability of our results to other clinical settings. *Sixth*, the small sample size made it difficult to find significant differences between subgroups, though non-significant trends could be highlighted. *Finally*, we planned to assess the use of bile acid resins, coenzyme Q10 niacin and olbetam, but they were not taken by any of the patients included in this study either.

While this study underlines the frequency of adverse effects, as well as the predominance of musculoskeletal symptoms caused by statins and other lipid-lowering drugs, it also shows that management is possible. Symptom intensity can be lowered through a number of strategies, such as a change of dosage, frequency of administration, daily timing, or active substance. As the benefit in terms of alleviation of symptoms compared to terminating treatment altogether does not show a significant difference (p=0.68) in this study, clinicians are encouraged to find a strategy tolerated by the patient. This is expected to raise treatment compliance and promotes the established benefits of lipid-lowering drugs in reducing the risk of CVD. Further research is required to compare different strategies and find ideal procedures for specific patient groups.

## Data Availability

The data underlying this article cannot be shared publicly because of patient's confidentiality issue.

## Acknowledgments

The authors would like to thank Prof. Nelly Pitteloud, Prof. Thierry Buclin and Dr. Haithem Chtioui, Lausanne University Hospital, Lausanne, Switzerland for their feedback on a previous version of the manuscript, as well as the patients and clinicians working at the Service of Endocrinology, Diabetes and Metabolism at Lausanne University Hospital, Lausanne, Switzerland.

---

**Supplementary Table 1.**
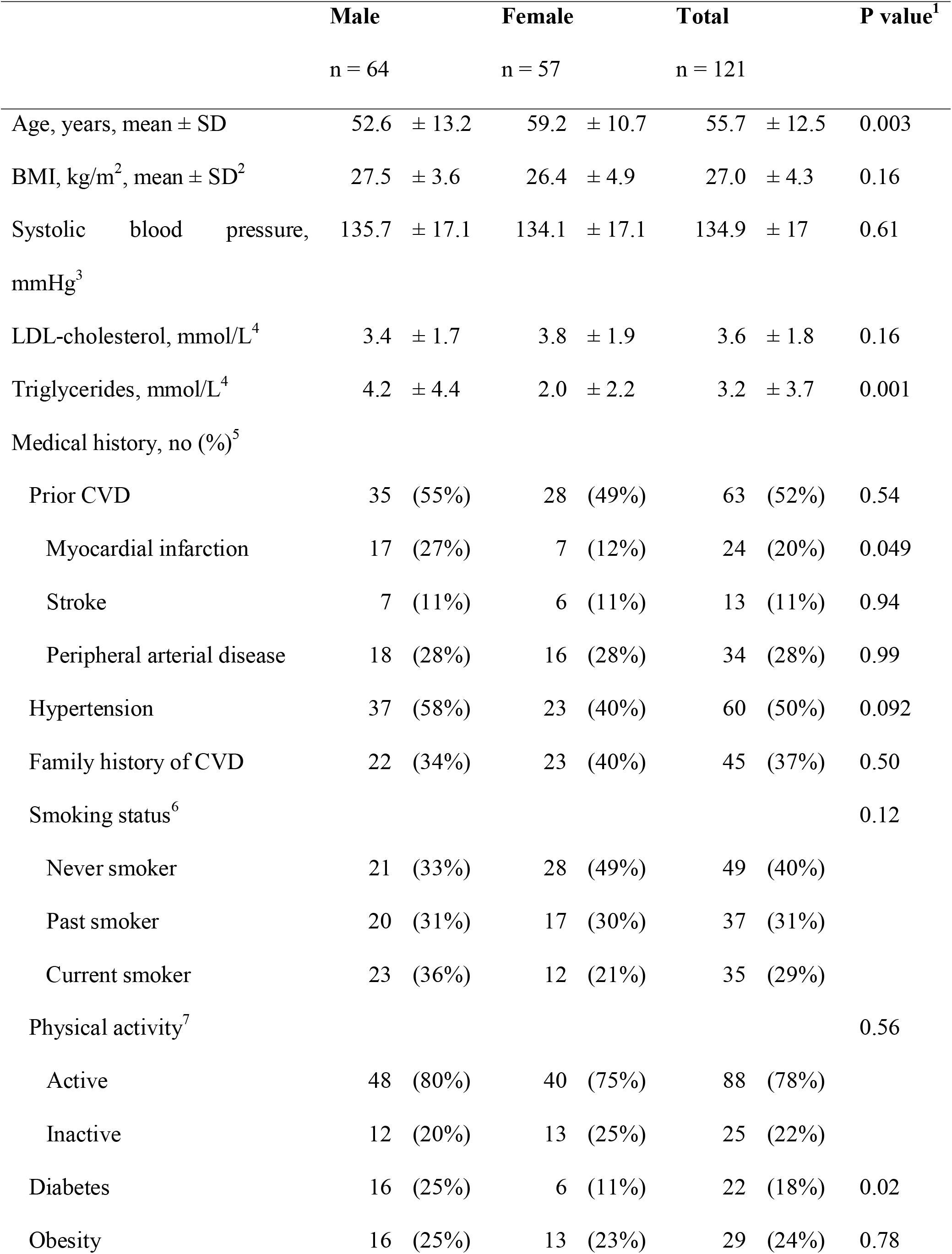
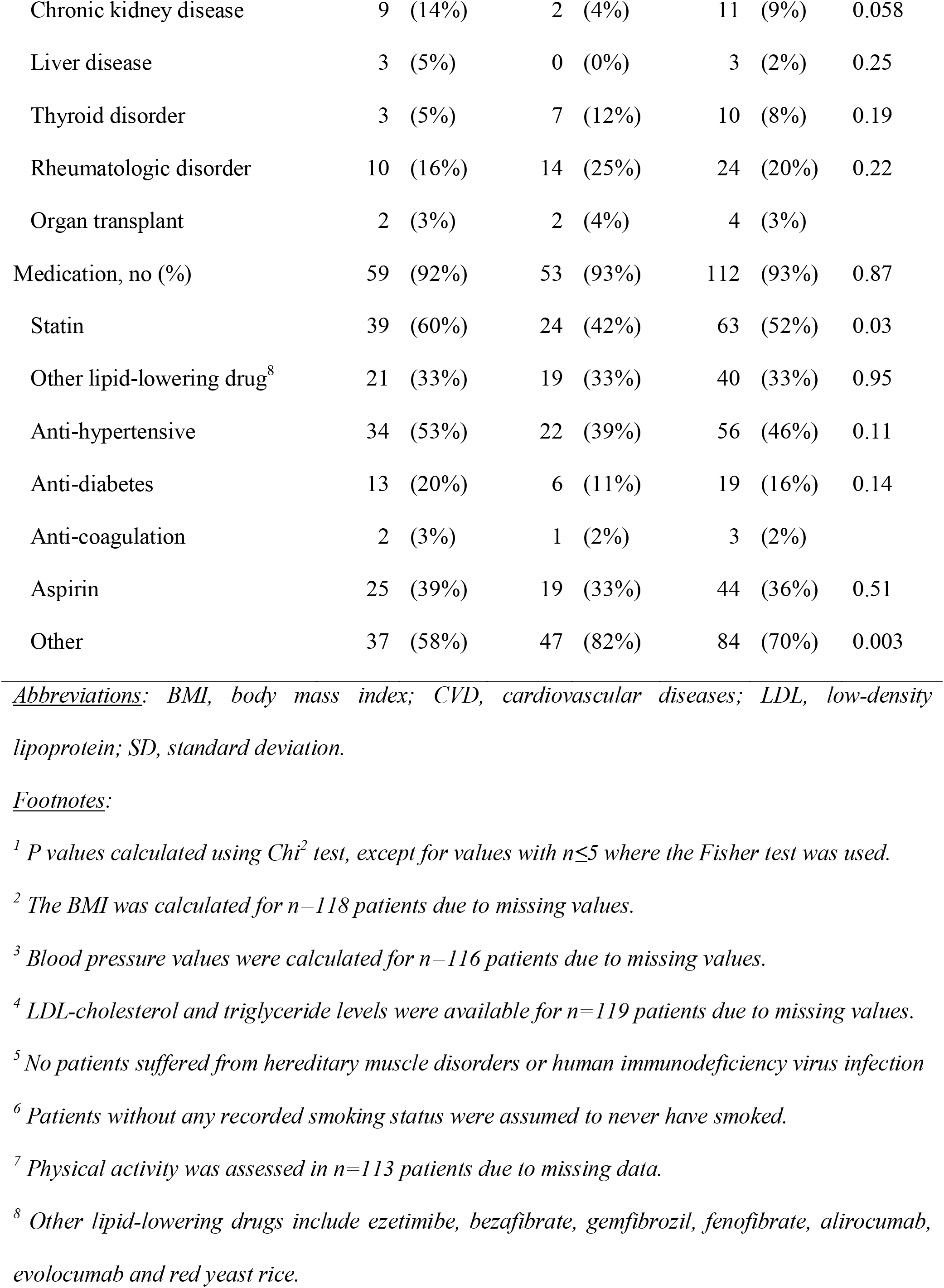
Clinical characteristics of 121 patients followed at lipid clinic, *details of Table 1*.

